# Rapid detection of SARS CoV-2 N501Y mutation in clinical samples

**DOI:** 10.1101/2021.04.17.21255656

**Authors:** Sirwan M.A. Al-Jaf, Sherko Subhan Niranji

**Affiliations:** Department of Biology, College of Education, University of Garmian, Kurdistan Region, Iraq; Coronavirus Research and Identification Lab., University of Garmian, Kurdistan Region, Iraq

**Keywords:** SARS CoV-2, Variant, N501Y, Mutation, Method, PCR

## Abstract

Severe acute respiratory syndrome coronavirus-2 (SARS CoV-2) variants poses major threats in increasing infectivity, transmission, mortality of Coronavirus Disease 2019 (Covid-19). Additionally, SARS CoV-2 variants resist antibody neutralizations or may abolish vaccine efficacies. Researches to develop economical and fast methods will support the developing or poor countries to challenge the Covid-19 pandemic via tracking common mutations that may help to deploy the vaccination programs and control the virus. Current study has developed a novel low-cost rapid technique, exploiting real time PCR probes and conventional PCR specific primers, to identify N501Y mutation, which was independently emerged in the UK, South African and Brazilian variants. Currently, these variants tend to spread to all over the world and seem to be more infectious, transmissible and fatal. This study helps tracking the N501Y mutation for understanding its clinical and epidemiological characteristics, in those countries where sequencing facilities are lacking or expensive. Further study should focus on other common mutations in the variants of concerns of SARS CoV-2.

## 1. Introduction

Several variants of SARS CoV-2 has been emerged following approximately a year of declaring the virus being pandemic by WHO (WHO, 2021). The variants, which usually emerge as accumulations of mutations that impact the survival fitness of the virus, transmissibility, infectivity, severity and fatality, are considered as variants of concern (VOC)(CD, 2021). Currently, the UK, South African and Brazilian variants have been three of the most common VOC that acquired a mutation called N501Y (Long et al., 2021), in which the wildtype nucleotide Adenine A23063 (Asparagine N501) in the spike protein of SARS CoV-2 was mutated to Thymine T23063 (Tyrosine Y501). This mutation, which is accompanied by several other mutations, happens in the spike protein and other proteins of SARS CoV-2 and collectively known as UK variant (N501Y V1, B.1.1.7, or VOC 202012/01) in the UK; South African variant (N501Y V2 or B.1.351) in South Africa and Brazil variant (N501Y V3, B.1.1.28 or P1) in Brazil.

Mutations occurred in the receptor binding domain (RBD) of SARS CoV-2 spike protein influence on the virus’s susceptibility to antibody neutralizations. Vaccines have exhibited variable potencies against the emerging VOC including the UK, South African and Brazilian variants. Sera of humans vaccinated with Moderna (mRNA-1273) or Pfizer (BNT162b2) show evasions from antibody neutralizations against the pseudoviruses that mimic SARS CoV-2 carrying either N501Y V2 or N501Y V3 variants (Garcia-Beltran et al., 2021; Hoffmann et al., 2021). The UK variant (N501Y V1) has been shown to be less susceptible to neutralizations by monoclonal antibodies but it showed susceptible to polyclonal antibodies taken from convalescent sera of humans previously infected or vaccinated with the wildtype strain (Rees-Spear et al., 2021). However, sera taken from Pfizer vaccinated persons show neutralization activities against both wildtype and mutant variants of the N501Y mutation (Shi et al., 2021). These studies indicate that the emerging variants have variable effects on antibody neutralizations and vaccine efficiencies. Nevertheless, the slight decline in the antibody neutralization in sera of the host population against the SARS CoV-2 variants (e.g., B.1.1.7 and B.1.351) may threaten protections of humans immunized with the SARS CoV-2 wildtype vaccines (Altmann et al., 2021). In addition, recent researches have linked the increased transmissibility, infectivity, mortality of SARS CoV-2 to the B.1.1.17 variant in the UK (Challen et al., 2021; Davies et al., 2021). These encourage the need for developing a method to explore N501Y mutation exploiting large number of samples clinically collected in patients infected with SARS CoV-2.

Rapid investigations of the N501Y mutation in clinical samples help the governments to take quick actions imposing emergency lockdowns against those emerging variants in both current or future local outbreaks. The prompt molecular techniques assist epidemiologists follow the distributions of the variants among populations in the past, current and future. Accordingly, vaccination programmes can be implemented for controlling further spread of the virus. Indeed, clinicians and immunologists can rely on the epidemiological and molecular biological data to understand the clinical and immunological aspects of the virus. This research aims to identify SASRS CoV-2 N501Y mutation in clinical samples using both primer-probe based real time reverse transcriptase polymerase chain reaction (rRT PCR) and endpoint PCR specific primers.

## 2. Material and Methods

### 2.1. Clinical samples

SARS CoV-2 positive nasopharyngeal samples (n=60) collected in VTM, were preserved in - 86°C freezer at Coronavirus Research and Identification Lab, University of Garmian, Kurdistan Region, Iraq. Consent forms were filled by the participants and the study was approved by an ethical committee at the Department of Biology, University of Garmian that follow the rules adhered to the Declaration of Helsinki for human and animal research.

### 2.2. Primer and probes

The Wuhan SARS CoV-2 whole genome sequence (GenBank: <u>MN908947.3</u>) was exploited to locate the primer and probes, which their melting temperatures, GC contents and amplification sizes were also inspected using online tools provided by NCBI and IDT websites. For the probe based rRT PCR, a pair of primers (for amplifying both sides of the N501Y mutation) and a pair of Taqman probes (each for identifying the wildtype N501 and mutant Y501) were designed (Macrogen, Korea). The probe for identifying the wildtype N501 was labelled with FAM dye while the probe for the mutant Y501 was linked with HEX dye. For the specific primer based endpoint PCR, one forward (F) primer and two reverse (R) primers were designed. One of the R primers was made for identifying the wildtype (N501 A mis R) and other for identifying the mutant (Y501 T mis R). Primers and probes were diluted to 10 pmole (10μM). Table 1 and 2 show all primers and probes used in this study. The locations of the primers and probes in SARS CoV-2 genome sequences was shown in Figure 1 A and the specific primers for the endpoint PCR was shown in Figure 1B.

**Table 1:**
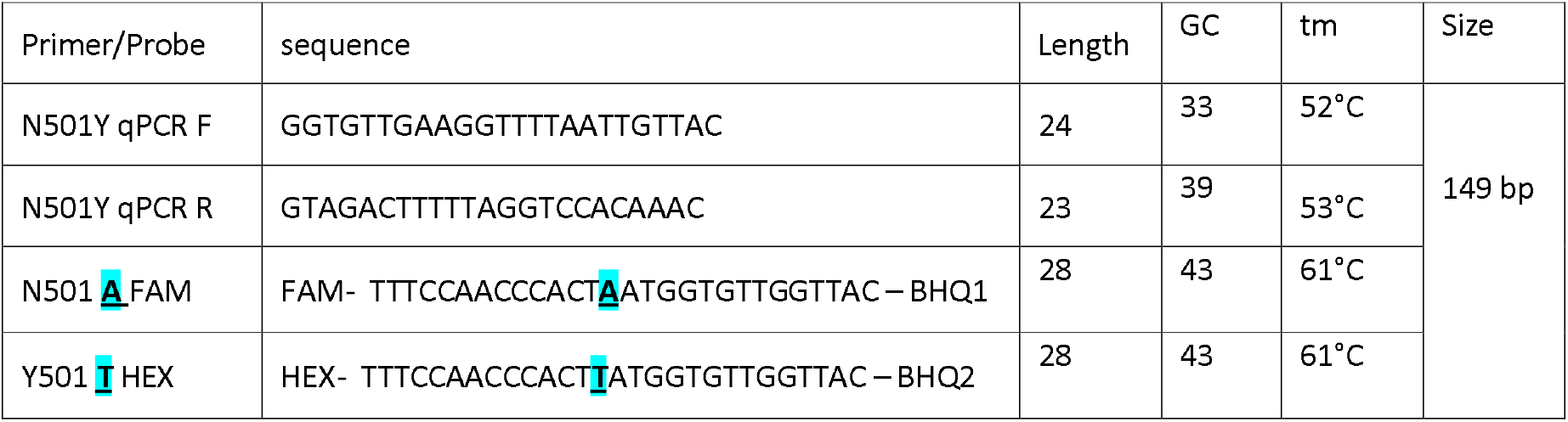
Primers and probes used for the rRT PCR. Underlined **<u>A</u>** and **<u>T</u>** in the FAM and HEX probes represent the nucleotides of the wildtype A23063 and mutant T23063 of the N501Y (A23063T) mutation.

**Table 2:**
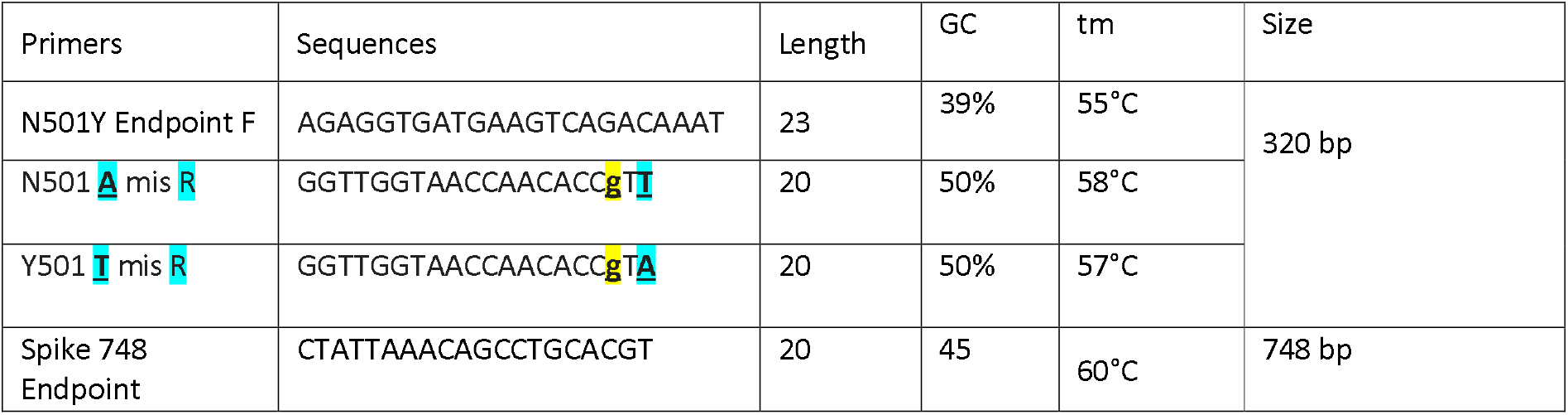
Primers used for the Endpoint PCR. The underlined **<u>T</u>** and **<u>A</u>** represent the reverse nucleotides of the wildtype A23063 and mutant T23063 of the N501Y (A23063T) mutation. The lower case g represents the mismatch made during designing the primers to reduce non-specific bindings.

**Figure 1:**
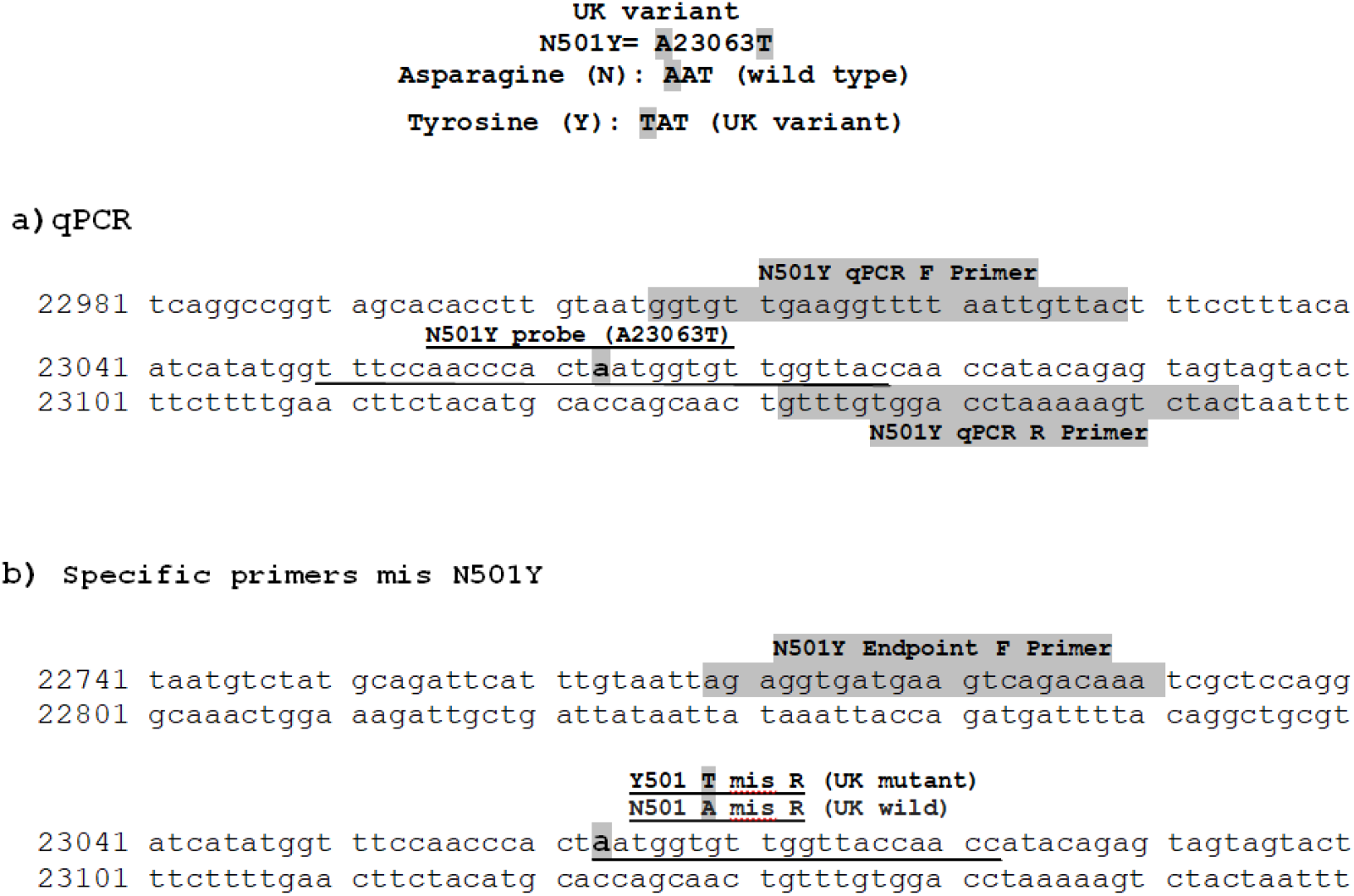
The positions of primers and probes used for rRT PCR (A) and Endpoint specific primers (B).

### 2.3. Identifications of SARS CoV-2

RNA extractions were performed for the VTM-stored clinical samples using automated RNA extraction machine and kits as recommended by the manufacturer (Nucleic Acid Extraction System BNP32, Biobase, China). The RNA samples were identified as SARS CoV-2 using either “Coronavirus COVID-19 genesig^®^ Real-Time PCR assay” (Primer design, UK) or “MutaPLEX^®^ Coronavirus real time RT-PCR kit” (immundiagnostik, Germany). As recommended by the manufacturer’s instructions, the RNA samples were kept at a -80°C freezer until the identification of the mutation was conducted.

### 2.4. Identifications of the N501Y mutation

#### 2.4.1. rRT PCR for identifications of N501Y

A mixture containing 8 μl of one step rRT PCR master mix (Addprobe, AddBio, South Korea), 0.5 μl of N501Y qPCR F primer, 0.5 μl of N501Y qPCR R primer, 0.5 μl of N501 A FAM probe, 0.5 μl of Y501 T HEX probe, and 5 μl of the viral RNA were run using CFX Connect Real-Time PCR Detection System (Bio-Rad, Germany). The rRT PCR reactions were performed as follow: reverse transcription at 50°C for 20 minutes, initial denaturation at 95°C for 10 minutes, followed by 50 cycles of denaturation at 95°C for 10 seconds and annealing at 65°C for 60 seconds.

#### 2.4.2. Endpoint PCR for identifications of N501Y

A mixture of 8 μl of one step rRT master mix (Addscript, Addbio, South Korea), 0.5 μl of N501Y Endpoint F primer, 0.5 μl of either “N501 A mis R” or “Y501 T mis R” (in a separate reaction), and 5 μl of RNA were performed as follow: reverse transcription at 50°C for 20 minutes, initial denaturation at 96°C for 10 minutes, followed by 36 cycles of denaturation at 95°C for 15 seconds, annealing at 66°C for 45 seconds and extension 72°C for 30 seconds and then a final extension at 72°C for 5 minutes using conventional PCR Lightcycler (Eppendorf, Germany).

#### 2.4.3. DNA sequencings

Six sequences, 4 N501 wildtype and 2 Y501 mutant samples were amplified using the N501Y Endpoint F primer with Spike 748 R (Table 2), that amplify a 748 bp product size on agarose gel electrophoresis and sent for Sanger sequencing (Macrogen Co., Seoul, KR). This was to confirm the N501Y mutants and other variant of concerns present in the UK, South Africa and Brazil variants. The sequences were submitted to NCBI using Bankit (Benson et al., 2015).

## 3. Results and Discussions

Current study aimed to identify the N501Y mutation in clinical samples. The rRT PCR results showed that both probes (N501 A FAM and Y501 T HEX) work at 65°C. Of 60 samples, 3 RNA samples carried Y501 mutant and 57 RNA samples were the N501 wildtype. Figure 2 shows examples of the N501 wildtype and Y501 mutant samples indicated by FAM and HEX signals, respectively.

**Figure 2:**
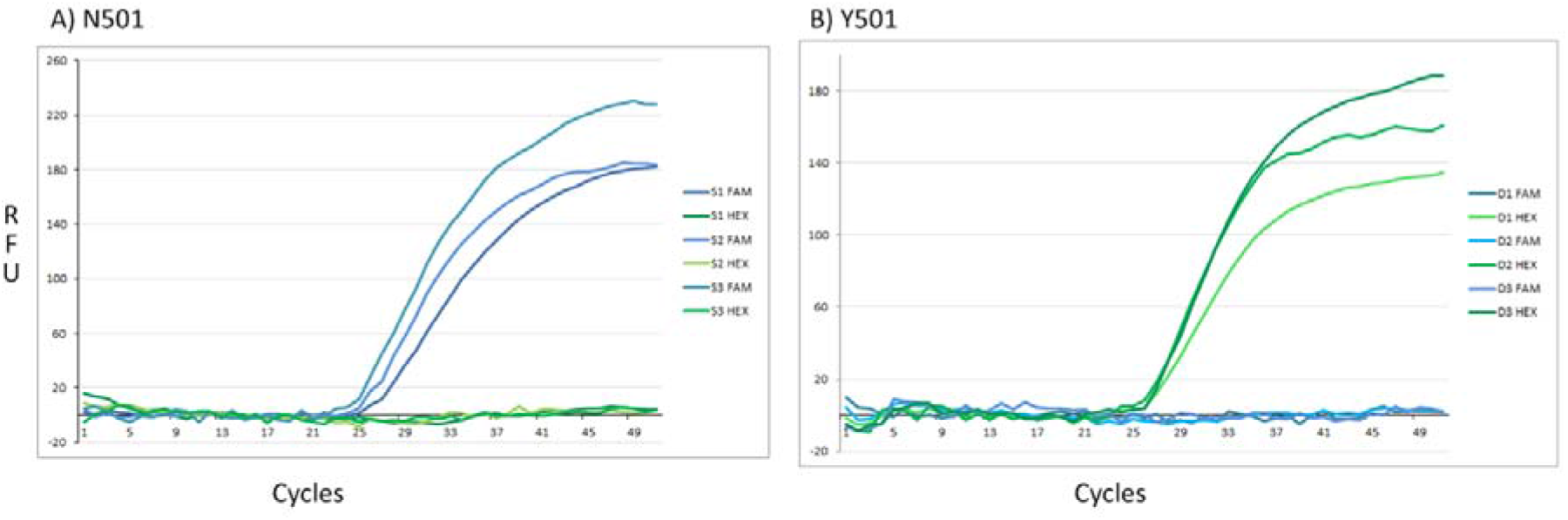
Taqman probe based rRT PCR. Three examples of the wildtype N501 (A) showed only FAM signals and the three samples of the mutant Y501 (B) showed only HEX signals.

The Endpoint PCR data confirmed the rRT PCR data as shown in Figure 3, which is an example of an N501 wildtype (Figure 3A), a Y501 mutant (Figure 3B). Two mutants and 4 wildtypes PCR products, amplified by conventional PCR amplifying 748 bp in the spike protein at the region of variants of concerns (Figure 3C), were confirmed using Sanger sequencings and the GenBanks (<u>MW897351</u>, <u>MW897352</u>, <u>MW897353</u>, <u>MW897354</u>, <u>MW897355</u>, and <u>MW897356</u>) were released to NCBI data base.

**Figure 3:**
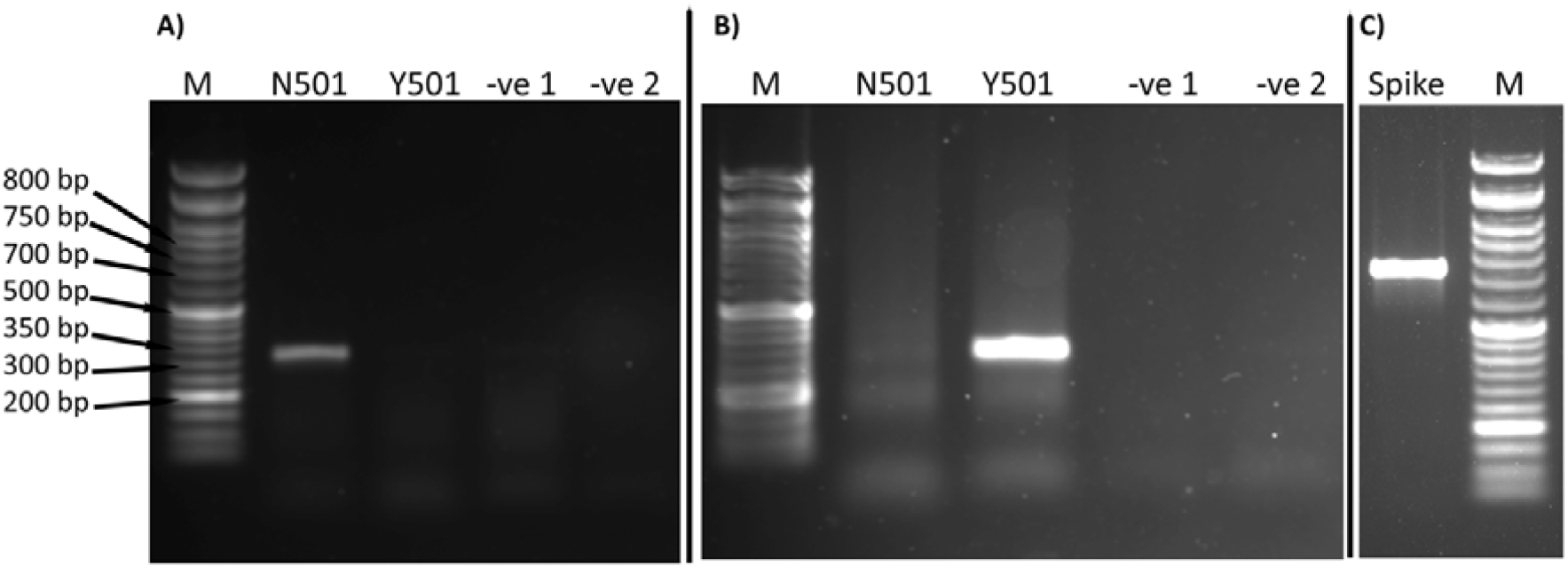
Agarose gel electrophoresis (1.5%). PCR products of reactions containing the specific primer of N501 A mis R wildtype (N501) or Y501 T mis R mutant (Y501) with the N501Y Endpoint F primer. -ve= contains only primers and master mix without RNA. M= Markers 50 bp. **Panel A:** an example of the wildtype N501. **Panel B:** an example of the mutant Y501. **Panel C:** an example of spike 748 primers sent for sequencing.

Our method can explore the N501Y mutation, which is present in the 3 variants of concern (VOC) that occurred in the UK (N501Y V1), South Africa (N501Y V2) and Brazil (N501Y V3). Additionally, the sequencing results showed that both D614G (A23403G) and A570D (C23271A), along with the N501Y (A23063T) mutations, also exist in Kurdistan region of Iraq. The D614G mutation has been dominant in almost all variants available in the world. However, the samples contained no other mutations, for example, E484K and K417N/T, which are present in both N501Y V2 and N501Y V3. This suggested that the N501Y mutations found in this study was originated from the N501Y V1 (UK variant), neither from the South African (N501Y V2) nor Brazilian (N501Y V3) variants. Frequent confirmations of SARS CoV-2 positive samples using approximately 750 bp of the spike protein by sequencings, as performed by the current study, will definitely help to identify new mutations. The spike 748 primers used in the current study can be used for sequencing other mutations in the RBD domain, such as Y453F in B.1.1.298 (the mink variant) and L452R in B.1.1.429 (California variant).

Up to our best knowledge, no N501Y mutation was published in Iraq and Kurdistan region. However, both governments have announced the presence of the UK variant in the country in late February 2021. Our mutant samples were taken on 23 March 2021. Epidemiological and phylogenetical analyses of samples, during February and March, is required to confirm the origin of the UK variant in Iraq, Kurdistan Region and their neighboring countries. Our rapid method will be advantageous over whole genome sequencings as the latter is expensive and time-consuming. Thus, the current developed technique is useful for surveying past and future clinical samples in developing countries as Iraq and its neighbors. In fact, this is important for tracking the N501Y mutation in poor countries or countries where no whole genome sequencing is available. Indeed, this method is also important for the clinicians and immunologists to conduct further research on the impact of this mutation on the transmissions, infectivity, severity and mortality that occurred in Covid-19 patients. Vaccination programs have been recently deployed in the Middle Eastern countries. Nonetheless, the developing and poor countries have not been sufficiently vaccinated or even vaccinations may have not reached in some countries in the world. Investigations of the common variants as the UK, South African and Brazilian variants are needed to apply any vaccines in any countries as some vaccines may not work against some variants.

During writing of this manuscript, a rapid method for screening N501Y mutations has been developed by Banadaa *et al* using melting curve analysis and sloppy molecular beacon probes (Banada et al., 2021). The limitation of the current work was that we can only explore the N501 mutation. Development of this method is required to identify other mutations such as K417N mutation, which co-occurred with N501Y in the South African B.1.351 variant, and K417T mutation, which is present in the Brazil variant. However, our sequencings found A570D and D614G mutations as well. Absence of K417N/T but presence of A570D in our sequences, suggested that the N501Y mutant samples are potentially the UK B.1.1.7 variant, not the South Africa or Brazil variants. Further study to find other common mutations such as K417N/T, E484K, P681H and Del69/70 residues of the spike protein should be taken into considerations.

## Conclusions

This study developed a novel method to discover N501Y mutation which is present in the most common variants of concerns including the UK, South African and Brazilian variants. This method can be applied to clinical samples all over the world since it is more inexpensive than while genome sequencings for tracking the mutation.

## Data Availability

). Two mutants and 4 wildtypes PCR products, amplified by conventional PCR amplifying 748 bp in the spike protein at the region of variants of concerns (Figure 3C), were confirmed using Sanger sequencings and the GenBanks (MW897351, MW897352, MW897353, MW897354, MW897355, and MW897356) were released to NCBI data base.

https://www.ncbi.nlm.nih.gov/nuccore/MW897351

https://www.ncbi.nlm.nih.gov/nuccore/MW897352

https://www.ncbi.nlm.nih.gov/nuccore/MW897353

https://www.ncbi.nlm.nih.gov/nuccore/MW897354

https://www.ncbi.nlm.nih.gov/nuccore/MW897355

https://www.ncbi.nlm.nih.gov/nuccore/MW897356

## Acknowledgement

We would like to thank Sheikh Hassan Al-Talabani for his great support in building our new laboratory, Coronavirus Research, and Identification Lab., at the time of Covid-19 pandemic. We are grateful to all patients for giving their samples.

## References

Altmann, D.M., Boyton, R.J., Beale, R., 2021. Immunity to SARS-CoV-2 variants of concern. Science (80-.). 371, 1103 LP – 1104. https://doi.org/10.1126/science.abg7404

Banada, P., Green, R., Banik, S., Chopoorian, A., Streck, D., Jones, R., Chakravorty, S., Alland, D., 2021. A Simple RT-PCR Melting temperature Assay to Rapidly Screen for Widely Circulating SARS-CoV-2 Variants. medRxiv Prepr. Serv. Heal. Sci. 2021.03.05.21252709. https://doi.org/10.1101/2021.03.05.21252709

Benson, D.A., Clark, K., Karsch-Mizrachi, I., Lipman, D.J., Ostell, J., Sayers, E.W., 2015. GenBank. Nucleic Acids Res. 43, D30–D35. https://doi.org/10.1093/nar/gku1216

CD, 2021. SARS-CoV-2 Variant Classifications and Definitions [WWW Document].

Challen, R., Brooks-Pollock, E., Read, J.M., Dyson, L., Tsaneva-Atanasova, K., Danon, L., 2021. Risk of mortality in patients infected with SARS-CoV-2 variant of concern 202012/1: matched cohort study. BMJ 372. https://doi.org/10.1136/bmj.n579

Davies, N.G., Jarvis, C.I., van Zandvoort, K., Clifford, S., Sun, F.Y., Funk, S., Medley, G., Jafari, Y., Meakin, S.R., Lowe, R., Quaife, M., Waterlow, N.R., Eggo, R.M., Lei, J., Koltai, M., Krauer, F., Tully, D.C., Munday, J.D., Showering, A., Foss, A.M., Prem, K., Flasche, S., Kucharski, A.J., Abbott, S., Quilty, B.J., Jombart, T., Rosello, A., Knight, G.M., Jit, M., Liu, Y., Williams, J., Hellewell, J., O’Reilly, K., Chan, Y.-W.D., Russell, T.W., Procter, S.R., Endo, A., Nightingale, E.S., Bosse, N.I., Villabona-Arenas, C.J., Sandmann, F.G., Gimma, A., Abbas, K., Waites, W., Atkins, K.E., Barnard, R.C., Klepac, P., Gibbs, H.P., Pearson, C.A.B., Brady, O., Edmunds, W.J., Jewell, N.P., Diaz-Ordaz, K., Keogh, R.H., Group, C.C.-19 W., 2021. Increased mortality in community-tested cases of SARS-CoV-2 lineage B.1.1.7. Nature. https://doi.org/10.1038/s41586-021-03426-1

Garcia-Beltran, W.F., Lam, E.C., St Denis, K., Nitido, A.D., Garcia, Z.H., Hauser, B.M., Feldman, J., Pavlovic, M.N., Gregory, D.J., Poznansky, M.C., Sigal, A., Schmidt, A.G., Iafrate, A.J., Naranbhai, V., Balazs, A.B., 2021. Multiple SARS-CoV-2 variants escape neutralization by vaccine-induced humoral immunity. Cell S0092-8674(21)00298–1. https://doi.org/10.1016/j.cell.2021.03.013

Hoffmann, M., Arora, P., Groß, R., Seidel, A., Hörnich, B.F., Hahn, A.S., Krüger, N., Graichen, L., Hofmann-Winkler, H., Kempf, A., Winkler, M.S., Schulz, S., Jäck, H.-M., Jahrsdörfer, B., Schrezenmeier, H., Müller, M., Kleger, A., Münch, J., Pöhlmann, S., 2021. SARS-CoV-2 variants B.1.351 and P.1 escape from neutralizing antibodies. Cell. https://doi.org/10.1016/j.cell.2021.03.036

Long, S.W., Olsen, R.J., Christensen, P.A., Subedi, S., Olson, R., Davis, J.J., Saavedra, M.O., Yerramilli, P., Pruitt, L., Reppond, K., Shyer, M.N., Cambric, J., Finkelstein, I.J., Gollihar, J., Musser, J.M., 2021. Sequence Analysis of 20,453 Severe Acute Respiratory Syndrome Coronavirus 2 Genomes from the Houston Metropolitan Area Identifies the Emergence and Widespread Distribution of Multiple Isolates of All Major Variants of Concern. Am. J. Pathol. https://doi.org/10.1016/j.ajpath.2021.03.004

Rees-Spear, C., Muir, L., Griffith, S.A., Heaney, J., Aldon, Y., Snitselaar, J.L., Thomas, P., Graham, C., Seow, J., Lee, N., Rosa, A., Roustan, C., Houlihan, C.F., Sanders, R.W., Gupta, R.K., Cherepanov, P., Stauss, H.J., Nastouli, E., Doores, K.J., van Gils, M.J., McCoy, L.E., 2021. The effect of spike mutations on SARS-CoV-2 neutralization. Cell Rep. 34, 108890. https://doi.org/10.1016/j.celrep.2021.108890

Shi, P.-Y., Xie, X., Zou, J., Fontes-Garfias, C., Xia, H., Swanson, K., Cutler, M., Cooper, D., Menachery, V., Weaver, S., Dormitzer, P., 2021. Neutralization of N501Y mutant SARS-CoV-2 by BNT162b2 vaccine-elicited sera. Res. Sq. https://doi.org/10.21203/rs.3.rs-143532/v1

WHO, 2021. SARS-CoV-2 Variants [WWW Document]. URL https://www.who.int/csr/don/31-december-2020-sars-cov2-variants/en/ (Accessed 4.11.21).

